# The usefulness of a quantitative olfactory test for the detection of COVID-19

**DOI:** 10.1101/2021.01.20.21250173

**Authors:** Marcos A Lessa, Stella M Cotta-Pereira, Frederico A Ferreira, Therezinha Marta P P Castiñeiras, Rafael M Galliez, Débora S Faffe, Isabela de C Leitão, Diana Mariani, Erica R Nascimento, Flávia S Lessa, Isabela Brasil Succi, Carlos A Pedreira

## Abstract

**Background:** During the COVID-19 pandemic, olfactory dysfunction (anosmia or hyposmia) has been reported by many patients and recognized as a prevalent and early symptom of infection. This finding has been associated with viral-induced olfactory neuron dysfunction rather than the nasal congestion typically found in cold- or flu-like states. In literature, the prevalence of anosmia varies from 15% to 85%, and the studies, in general, were based on the subjective evaluation of patients’ self-reports of loss of smell (yes or no question). In the present study, we quantitatively evaluated olfactory dysfunction and the prevalence of fever in symptomatic patients suspected of having COVID-19 using a scratch-and-sniff olfactory test and infrared temperature testing with RT-PCR as the gold-standard comparator method to diagnose COVID-19 infection.

**Methods:** Outpatients had their forehead temperature checked with an infrared non-contact thermometer (temperature guns). After that, they received two olfactory smell identification test (SIT) cards (u-Smell-it™; CT, USA) that each had 5 scent windows and were asked to scratch with a pencil and sniff each of the 10 small circles containing the microencapsulated fragrances and mark the best option on a response card. Nasopharyngeal swabs were then collected for Reverse Transcriptase-Polymerase Chain Reaction (RT-PCR) to determine if the patients were positive or negative for COVID-19 infection. We considered the number of ‘hits’ (correct answers) ≤ 5 as positive for loss of smell (LOS) in the olfactory test; ≥ 6 hits was considered negative for LOS (i.e. normal olfactory function). All data were analyzed using Excel and Matlab software.

**Results:** In the present study, 165 patients were eligible for the olfactory test and nasopharyngeal swab collection RT-PCR. Five patients were excluded because of inconclusive PCR results (n=2) and missing data (n=3). A total of 160 patients completed all the protocols. The RT-PCR positivity rate for COVID-19 was 27.5% (n=44), and PCR+ patients scored significantly worse in the olfactory test (5.5±3.5) compared to RT-PCR-patients (8.2±1.8, p<0.001). 0/44 PCR+ patients presented with a fever (≥37.8°C). In contrast an olfactory SIT had a specificity of 94.8% (95% CI, 89.1 – 98.1), sensitivity of 47.7% (95% CI, 32.7 – 63.3), accuracy of 0.82 (95% CI, 0.75 – 0.87), positive predictive value of 77.8% (95% CI, 59.6 – 88.8), negative predictive value of 82.7% (85% CI, 78.7 – 86.7), and odds ratio of 16.7.

**Conclusion:** Our results suggest that temperature checking failed to detect COVID-19 infection, while an olfactory test may be useful to help identify COVID-19 infection in symptomatic patients.

## Introduction

The coronavirus disease 2019 (COVID-19) can cause severe injury and death and quickly became a pandemic as the transmission of the virus often occurs before or at the onset of symptoms. To further complicate matters, the symptoms associated with COVID-19 are variable and many can be mild and hard to objectively identify, or even not present at all (asymptomatic). Thus, challenges to science and medicine are to stop the rapid viral transmission and identify suspicious cases in the context of this wide range of symptoms. Among the clinical manifestations of COVID-19, olfactory dysfunction (OD), which consists of anosmia or hyposmia, is frequently reported by many patients, and studies, which are based mainly on patient surveys and have indicated that OD is a prevalent and early symptom of SARS-CoV-2 infection^1–4^. Distinct from the nasal congestion typically found in viral upper respiratory infections, covid-induced OD is associated with the virus’s presence in cells adjacent to the olfactory neurons^5^ and, which by a mechanism that is not fully elucidated, can cause alterations in the odor perception function of olfactory neurons^5^. The quick onset of anosmia is a highly suggestive symptom of the SARS CoV-2 viral infection, and, in many cases of COVID-19, OD and ageusia are the only presenting symptom^3,4,6^. However, OD, especially hyposmia, may be unnoticed unless formally and objectively tested with a measurable olfactory test.

There a wide range for the prevalence of OD in the literature, from 15% to 85%, and most of the data is from subjective evaluation of patients’ self-reports of loss of smell^1,7–9^. The use of an olfactory test for the diagnose of COVID-19 infection has been recently reported in different studies^7,9–13^. However, nearly all those studies only tested the OD in PCR positives patients. Thus, the test’ specificity and degree of association with the disease (full odds ratio) are unclear. Similarly, how quantitative olfactory tests compares to temperature testing, which is also a quantifiable symptom and is commonly used for COVID-19 screening, is unclear for both have not been performed on the same subjects.

Population testing is a crucial strategy for identifying and isolating suspicious cases efficiently, mainly because of the COVID-19 high infectivity. The principal tool to diagnosing COVID-19 is Reverse Transcriptase-Polymerase Chain Reaction (RT-PCR) and, to a lesser extent, antigen tests. While PCR is the gold standard and has excellent sensitivity and specificity, it is costly and requires sample collection by others, special handling, instrumentation, and analysis - often at distant locations that can cause significant delays. These issues pose challenges for very wide scale population testing and are more largely confined to testing a subpopulation of suspected infected people. There is a world need for an alternative, affordable and reliable, albeit not perfect, test to be used on a large scale to more easily identify infected individuals and block the transmission.

Here, we compared scratch-and-sniff style smell identification test (SIT) that has 5 odorants windows on a single card as a potentially quick, inexpensive, and easy pre-screen test of COVID-19 in symptomatic patents, with RT-PCR as the reference standard to investigate the feasibility of using a quantifiable olfactory test to help identify COVID-19 infection. Also, we examined the effectiveness of infrared forehead temperature screens in the same patient cohort.

## Methods

The study was carried out at the Center for COVID-19 diagnosis of the Federal University of Rio de Janeiro (UFRJ). A total of 165 individuals that attended the Center presenting mild cold-like symptoms were enrolled from June to August, 2020. Nasopharyngeal swabs were obtained from each participant and COVID-19 diagnosis was performed by RT-PCR using the CDC protocol, with primers and probes for N1 and N2 targets. Clinical and demographic data were self-reported by the patients. The study was approved by the local ethics committee from Clementino Fraga Filho University Hospital (CAAE: 30161620.0.0000.5257). Written informed consent was obtained from all participants. Inclusion criterium was symptomatic outpatients older than 18 years scheduled to collect nasopharyngeal swab for PCR test at UFRJ testing facility. Patients that had a rhinorrhea or nasal congestion were excluded and eligible volunteers had their forehead temperature checked with an infrared non-contact thermometer (temperature guns). After that, the volunteers received two ‘u-Smell-it™ ’ (Connecticut, USA) olfactory SIT cards (5 scents each) and were asked to scratch and sniff each of the 10 areas containing the microencapsulated fragrancies and mark the best choice of 5 options (4 scent choices and ‘no scent’) on a response card. In our protocol, three versions of the test were used. The cards #1414 and #1515 have the same scents, however they are in different order of presentation, and the card #1313 has different scents from #1414 and #1515. Supplemental Figures 1 and 2 present examples of card test and response card, respectively. For the first protocol, we used one card #1313 in combination with another card #1414 or #1515 to have 10 different smells presented to the patients. For the second protocol (reproducibility analysis), we used #1414 and #1515 cards to have 2 smells repeated in both cards. Supplemental Table 1 shows the scent options and the right response for all cards used in this study.

For tests with two olfactory cards (10 scents) the optimal cutoff to most accurately distinguish COVID-19 positive and negative patients, as determined by RT-PCR, was achieved using ≤ 5 number of correct responses (‘hits’) as scored as ‘positive’ for loss-of-smell (LOS); ≥ 6 hits was considered as negative for LOS. Figure 1 shows the sensitivity and specificity for different cutoffs and the ROC curve for the test using two cards.

**Figure 1.**
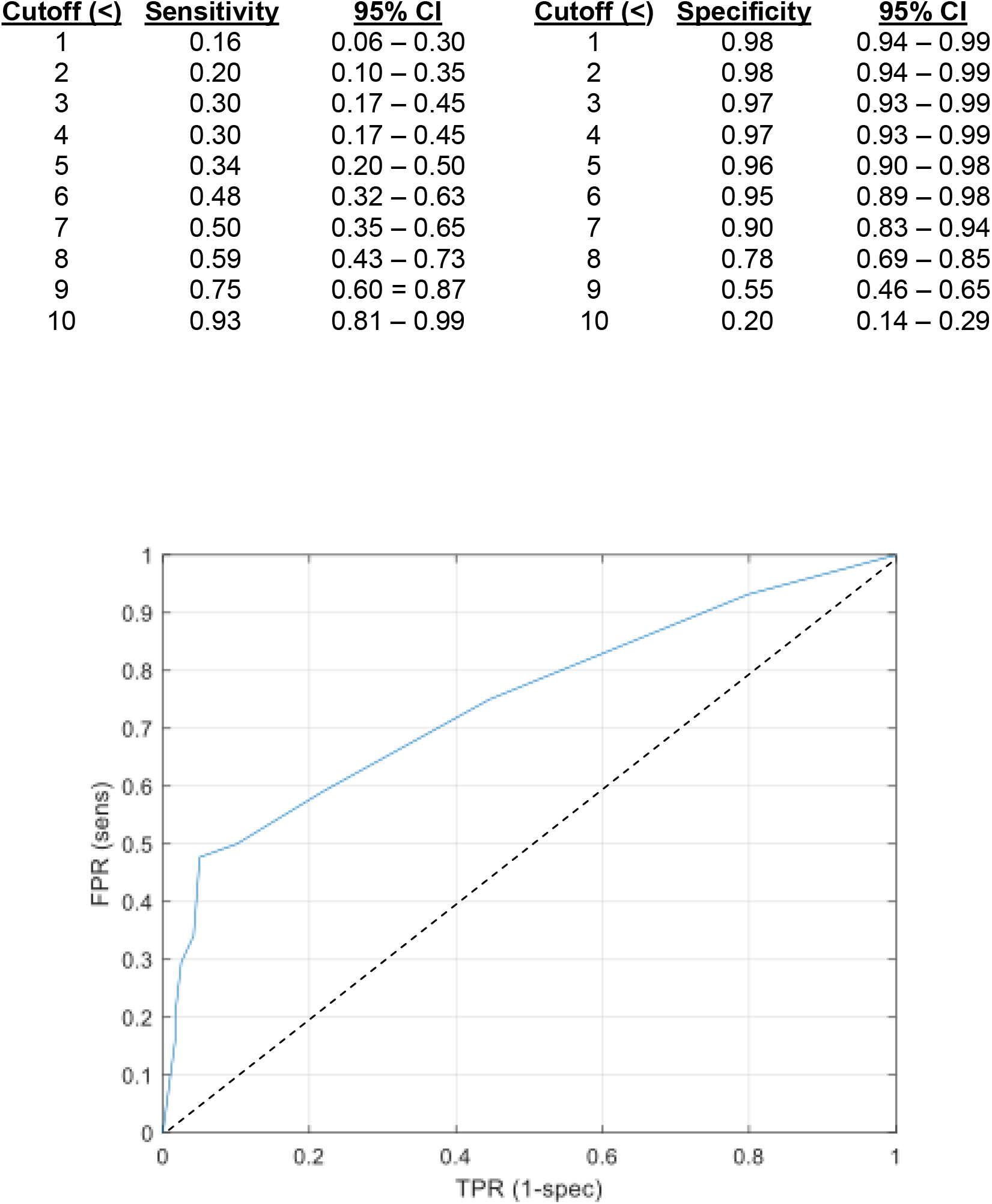
Sensitivity and specificity and ROC curve for U-smell-it™ used with two cards (10-scent).

To evaluate if the use of a single olfactory card (5 scents) presents similar results when comparing with the use of two olfactory cards (10 scents) we used a bootstrap statistical procedure (see below) to simulate the use of a single card and determine the statistical results. Figure 2 shows the sensitivity and specificity for different cutoffs after bootstrap statistical procedure and the ROC curve.

**Figure 2.**
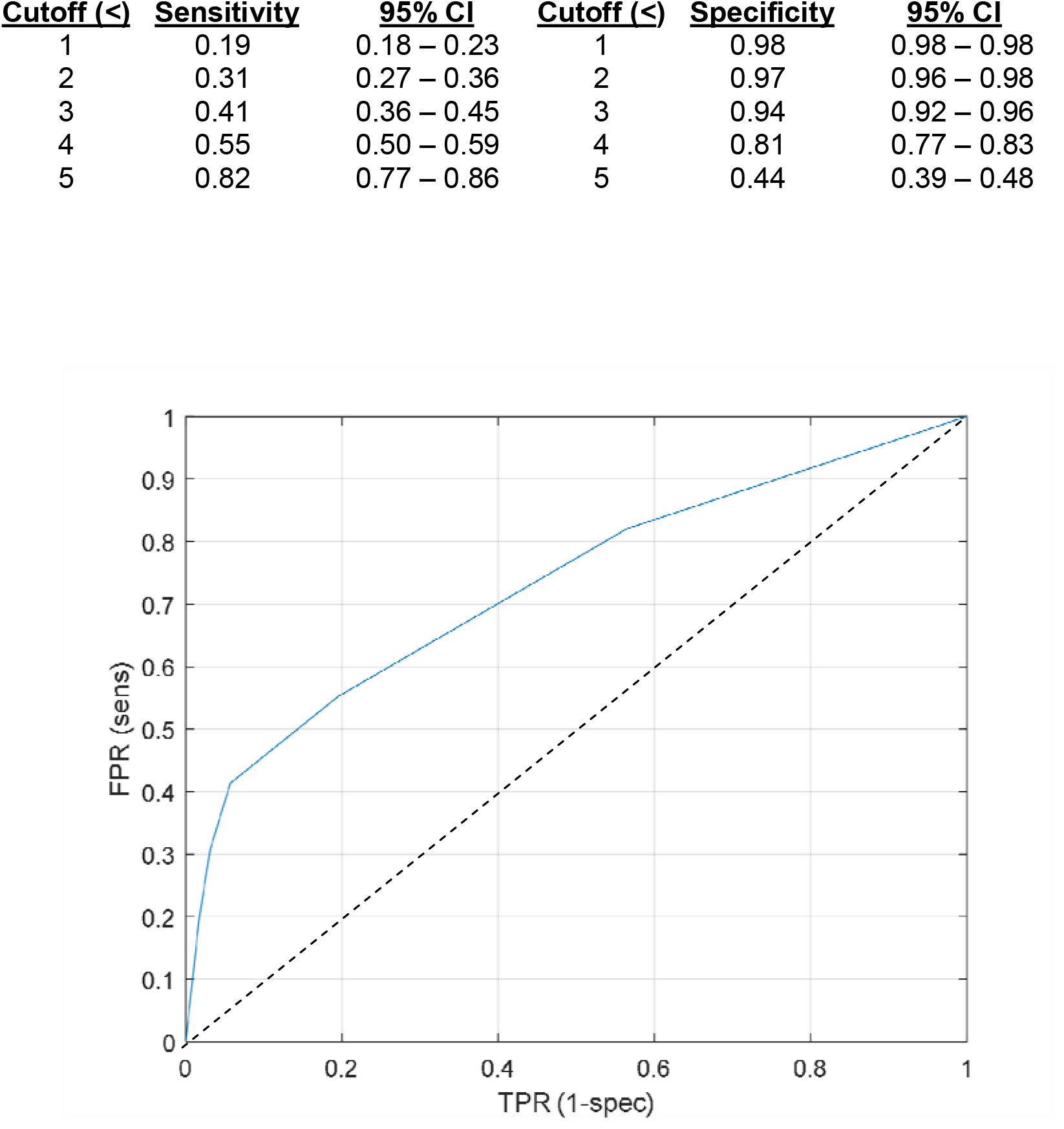
Sensitivity and specificity and ROC curve for U-smell-it™ used as a single card (5-scent).

### Statistical methodology

With the goal of testing for a positive association of the U-smell-it™ olfactory test with the RT-PCR based COVID-19 diagnosis, two binary variables were considered, one for the tested scheme and another for the gold/reference standard diagnosis. We built contingency tables and calculated sensitivity, specificity, positive and negative predictive values. We also applied the Chi square Pearson test^14,15^ to demonstrate that the proposed method does not produce an independent outcome in comparison with the gold standard diagnosis.

Bootstrap^16^ is a statistical procedure that creates a large quantity of simulated samples by resampling the dataset. The parameters are estimated by averaging the results of all runs. Here, we randomly sorted 5 out of 10 odorants for each patient and calculated the parameters after passing through the complete dataset. This procedure was repeated 10,000 times for the whole data set, generating 10,000 values for each parameter. Note that new random draws were performed for each patient in all the runs. At the end, the average of these 10,000 runs were used to estimate for the parameters and confidence intervals, as calculated through the percentiles 2.5 and 97.5.

Concerning reproducibility, a two-branch experiment was run with 5 odorants in each for the same group of individuals to evaluate divergencies between branches responses. The Wilcoxon signed rank test was used for the post-hoc analyses.

All data was statistically analyzed Matlab software (R2019b) and confirmed with Medcalc® software.

## Results

In the present study, 165 patients were eligible for the olfactory test and nasopharyngeal swab collection for PCR. Five patients were excluded because of an inconclusive PCR result (n=2) and missing data (n=3). A total of 160 patients completed all the full protocol. The PCR positivity rate for COVID-19 was 27.5% (n=44). Demographic data are shown in Table 1.

**Table 1.** Demographic data

|  |  | All patients<br>(n=160) | RT-PCR+ patients<br>(n=44) | RT-PCR– patients<br>(n=116) |
| --- | --- | --- | --- | --- |
| Age |  | 38.3±13 | 39.5±12.4 | 35.5±13.2 |
| Gender | M (n, %) | 54 (33.8) | 20 (45.4) | 34 (29.3) |
|  | F (n, %) | 106 (66.2) | 24 (54.6) | 82 (70.7) |
| Forehead temp (°C) |  | 36.4±0.2 | 36.4±0.2 | 36.34±0.2 |
| Self-report LOS (n, %) |  | 60 (37.5) | 28 (63.6) | 32 (27.5) |
| U-Smell-it mean score |  | 7.5±2.7 | 5.5±3.5* | 8.2±1.8 |

LOS, loss of smell.

In our study, a forehead temperature check did not identify any case of fever (0/165) as defined as temperature of 37.8°C or above. The frequency distribution of the temperatures is shown in Figure 3.

**Figure 3.**
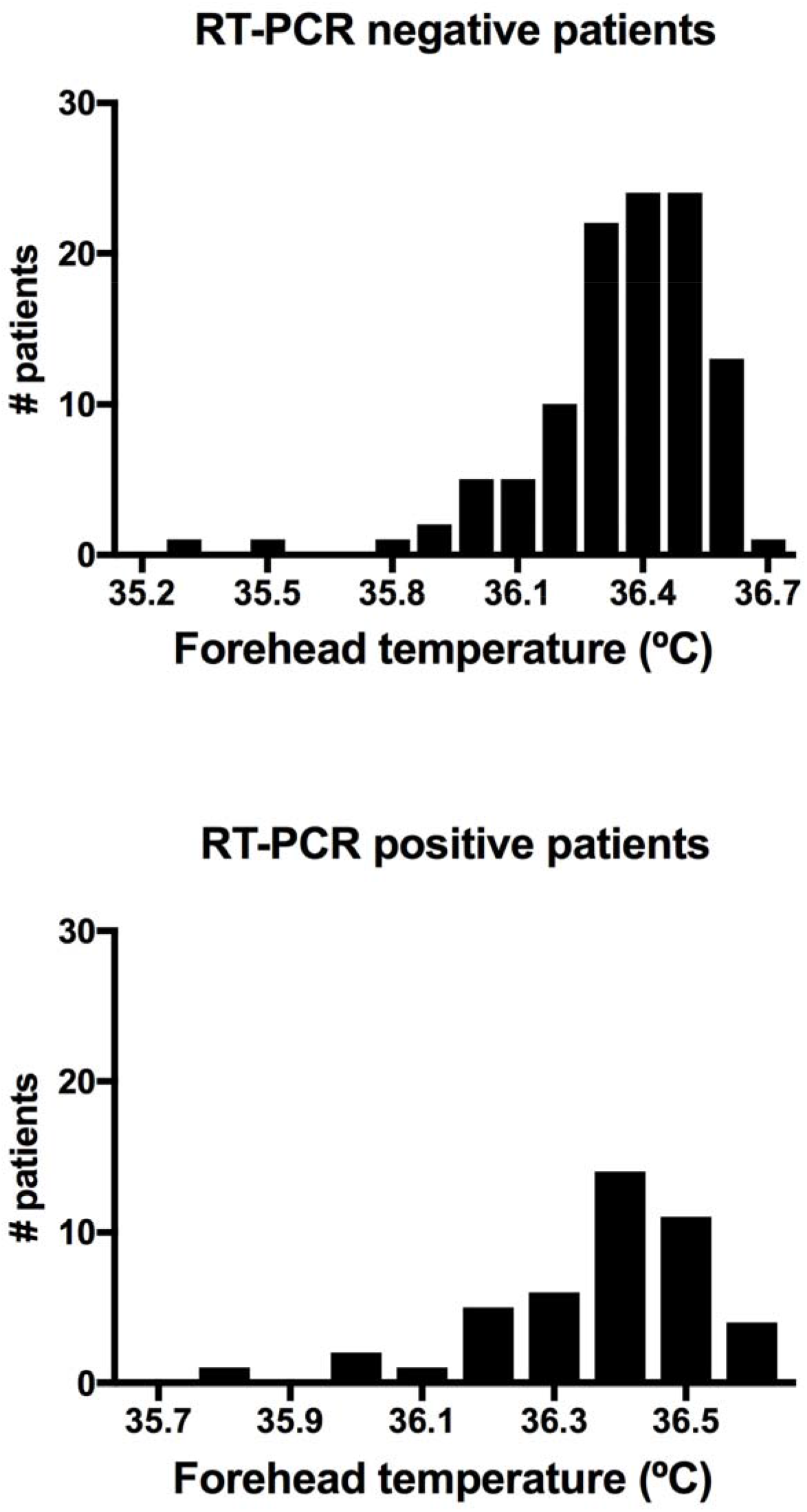
Frequency distribution of forehead temperature of patients according RT-PCR result.

The contingence table and the statistical analysis of the olfactory test performance using two u-Smell-it™cards (10 scents) when comparing with RT-PCR results for SARS-CoV-2 are shown in Tables 2 and 3, respectively. Olfactory testing showed a specificity of 94.8% (95% CI, 89.1 – 98.1), sensitivity of 47.7% (95% CI, 32.7 – 63.3), positive predictive value of 77.8% (95% CI, 59.6 – 88.8), negative predictive value of 82.7% (85% CI, 78.7 – 86.7), accuracy of 82% (95% CI, 75 – 87), and odds ratio of 16.7.

**Table 2.** U-smell-it™ olfactory test (two cards) vs RT-PCR

|  |  | RT-PCR |  |
| --- | --- | --- | --- |
|  |  | Positive | Negative |
| U-smell-it™ | Positive | 21 | 6 |
|  | Negative | 23 | 110 |

**Table 3.** Statistical findings of U-smell-it™ olfactory test (two cards) vs RT-PCR

|  | **Mean** | **95% IC** |
| --- | --- | --- |
| Sensitivity | 47.7% | 32.5 – 63.3% |
| Specificity | 94.8% | 89.1 – 98.1% |
| PPV | 77.8% | 59.6 – 88.8% |
| NPV | 82.7% | 78.7 – 86.7% |
| LR+ | 9.2 | 4.0 – 21/3 |
| LR- | 0.6 | 0.4 – 0.7 |
| OR | 16.7 | 6.1 – 46.1 |
| Accuracy | 82% | 75 – 87% |
| p-value | <0.0001 |  |
| Chi stat | 41.0 |  |

PPV, predictive positive value; NPV, negative predictive value; LP+, positive likelihood ratio; LR-, negative likelihood ratio; OR, odds ratio.

The contingence table and the statistical analysis of self-report of LOS performance when comparing with RT-PCR are shown in Tables 4 and 5, respectively. Although the sensitivity and PPV of self-report of LOS were higher than U-smell-it□ olfactory test, the specificity, NPV, and accuracy of a self-report were lower. Interestingly, using a RT-PCR test as a reference standard, the odds ratio for olfactory dysfunction as determined by a quantitative test (u-Smell-it™, 16.7) was 3.6-fold higher than the odds ratio of self-reported LOS (4.6).

**Table 4.** Self-report of loss of smell vs RT-PCR.

|  |  | PCR |  |
| --- | --- | --- | --- |
|  |  | Positive | Negative |
| Self-report LOS | Positive | 28 | 32 |
|  | Negative | 16 | 84 |

LOS, loss of smell.

**Table 5.** Statistical findings when comparing self-report of LOS to PCR.

|  | **Mean** | **95% IC** |
| --- | --- | --- |
| Sensitivity | 63.6% | 47.8 – 77.3% |
| Specificity | 72.4% | 37.1 – 55.3% |
| PPV | 46.0% | 59.6 – 88.8% |
| NPV | 84.3% | 78.2 – 89.0% |
| LR+ | 2.3 | 1.6 – 3.3 |
| LR- | 0.5 | 0.3 – 0.8 |
| OR | 4.6 | 2.2 – 9.6 |
| Accuracy | 70% | 62 – 77% |
| p-value | <0.0001 |  |
| Chi stat | 41.0 |  |

PPV, predictive positive value; NPV, negative predictive value; LP+, positive likelihood ratio; LR-, negative likelihood ratio; OR, odds ratio.

To evaluate the reproducibility of the quantitative olfactory test, another group of 66 patients was tested with two u-Smell-it™cards containing the same five scents, but were arranged in a different order. Forty-one out of 66 tested individuals produced exactly the same number of correct hits in both branches, 23 had a divergence of just 1 scent, and 2 patients mismatched 2 or more scents. Wilcoxon signed rank analysis produced a p value= 0.58, indicating that there is no evidence the ‘smells’ were perceived as diverse in the two branches of tests.

There were no side effects or complaints related to test participation of the subjects. Most patients took approximately 90 sec or less to complete the test with two olfactory cards (10 scents).

## Discussion

The present study showed that a large fraction of SARS-CoV-2 PCR+ patients had olfactory dysfunction, and this symptom, when detected by an olfactory test, was highly specific (95%). Our results demonstrated that the u-Smell-it™test’s odds ratio to detect COVID-19 infection was 3.6-fold higher than in self-reports, supporting that a quantitative test outperforms self-surveys. These findings were seen in the aggregate and after using a cutoff score of 3 on a 6 point scale (LOS = score 0, 1, 2 out of 5 maximum). The results indicate that a simple 5- or 10-window olfactory smell identification test can specifically differentiate, with only about 5% false positive, people infected by SARS-CoV-2 with 82% accuracy. Interestingly, a five-window test’s statistical performance metrics was similar to that of a 10-window test, compatible with the relative intense loss of smell in our study population. Dramatically, and in contrast to olfactory tests, temperature checking failed to detect any covid-19 infection. The study underscores the weakness and likely the limited usefulness of identification of COVID-19 individuals using temperature tests and infrared camera monitoring, which are used ubiquitously here in Brazil and many other countries to detect suspected cases. Our results show that among the 44 patients that were positives for SARS-CoV2, none had fever (as typically defined as a temperature over 38°C). Notably, fever is a non-specific symptom of viral infection, and because of that, the usefulness of temperature screening to identify suspicious cases has been called into question^17–20^. Nevertheless, body temperature checks are applied routinely as the primary screening test to identify individuals with fever in the entrance of many public places, such as schools, airports, hospitals, etc.

Of note, literature data show that only about 20% of people that have COVID-19 have a fever, and generally, this occurs very early on the course of the infection and has a short duration (under three days)^21^. Our SARS CoV-2 positive patients’ body temperature results were not significantly different from negative ones (Table 1 and Figure 1), and one possible explanation is that the outpatients observed in our study population may arrived after transiently having fever. It also shows the poor performance of temperature-based screens and further questions their usefulness as a screening test. Our study tested two different symptoms of COVID-19 infection (OD and temperature) and provided strong evidence that an olfactory test would significantly outperform a temperature test.

Another key finding of the study was that an olfactory test significantly outperformed self-reported LOS. Namely, to identify a patient with positive RT-PCR for COVID-19, the odds ratio for the self-reports was only 4.6, while using a real olfactory test, it was 16.7. This result suggests that patients with OD diagnosed using olfactory tests were more highly associated (3.6x) to test positive for COVID-19 than self-reports of LOS. This finding is consistent with other reports whereby an objective test outperformed non-objective and non-quantitative surveys^22^. Patients that failed to notice LOS included those with both anosmia and hyposmia.

Our results showed that the olfactory test specificity to detect OD in patients infected with COVID-19 was very high (∼95%), which is consistent with other reports^23^ including by the CDC^24^ indicating that “a new loss of taste or smell” is the single best indicator symptom of COVID-19, as based on the odds ratio. Although OD is acknowledged by many research teams and listed on the WHO website as a finding consistent with SARS-CoV-2 infection, little attention is given that OD is a better indicator of COVID-19 than other symptoms. Considering this symptom’s very high specificity, OD should be much better suited to rapid screening than the ubiquitous temperature test. The journal STAT news had proposed that, for the for screening of suspicious cases of COVID-19 infection, a smell test would be a better option than temperature tests^25^.

The sensitivity of the testing with U-smell-it™ (ranging from 48% in 10-scent test to 55% in 5-smell test) is lower than expected considering other reports that show OD detection via smell testing with a sensitivity of 76% (51–91%)^23^. Given that no patients had a fever in our population, and the average duration of loss of smell was reported to be around 7 to 8 days^11^, one possible interpretation is that some patients may have arrived after recovering from LOS. Consistent with this possibly no patients had an elevated temperature even above baseline. Secondly, from the self-reporting of loss of smell, the sensitivity was higher; however, with very poor specificity. Considering that the average duration of LOS is only about one week (note that a subset of patients (∼10%) can have a longer term loss) and that RT-PCR positivity may remain for three weeks or more, it is conceivable that, in our study, some patients were tested for OD after recovery; this is consistent with the finding indicating they had a recent LOS on the questionnaire but showing standard performance in an olfactory test. However, to properly address this would require longitudinal testing or other mechanisms to accurately identify disease staging, such as serology.

The study’s limitations include that we recruited only outpatient that came into the clinic for a diagnostic test at variable points of the disease, that as mentioned above we not ascertained, and we did not investigate asymptomatic patients. More, our cohort did not include children, pregnant, or the elderly, so our results may not be directly extrapolated to these groups of patients. It is anticipated that a short 5- or 10-odorant smell identification test, patients with minor hyposmia cases may be missed compared to a 40-odorant UP-SIT test^11,26^.

In conclusion, our study demonstrates that patients with positive SARS CoV-2 RT-PCR were highly associated with olfactory dysfunction, and 3.6-fold higher when tested with a short olfactory test than seen in self-reports. Taking together, these results suggest that quick olfactory tests may be useful to detect and COVID-19 infection in symptomatic patients.

## Supporting information

Supplemental_file_olfactory _test_paper

## Data Availability

All data from this paper will be available at the Brazilian Registry of Clinical Trials (ReBEC)

https://ensaiosclinicos.gov.br/welcome#menu

