## Supplemental_file_olfactory _test_paper for "The usefulness of a quantitative olfactory test for the detection of COVID-19"

Supplemental Figure 1. Example of a scratch-and-sniff (u-Smell-it™) olfactory test.

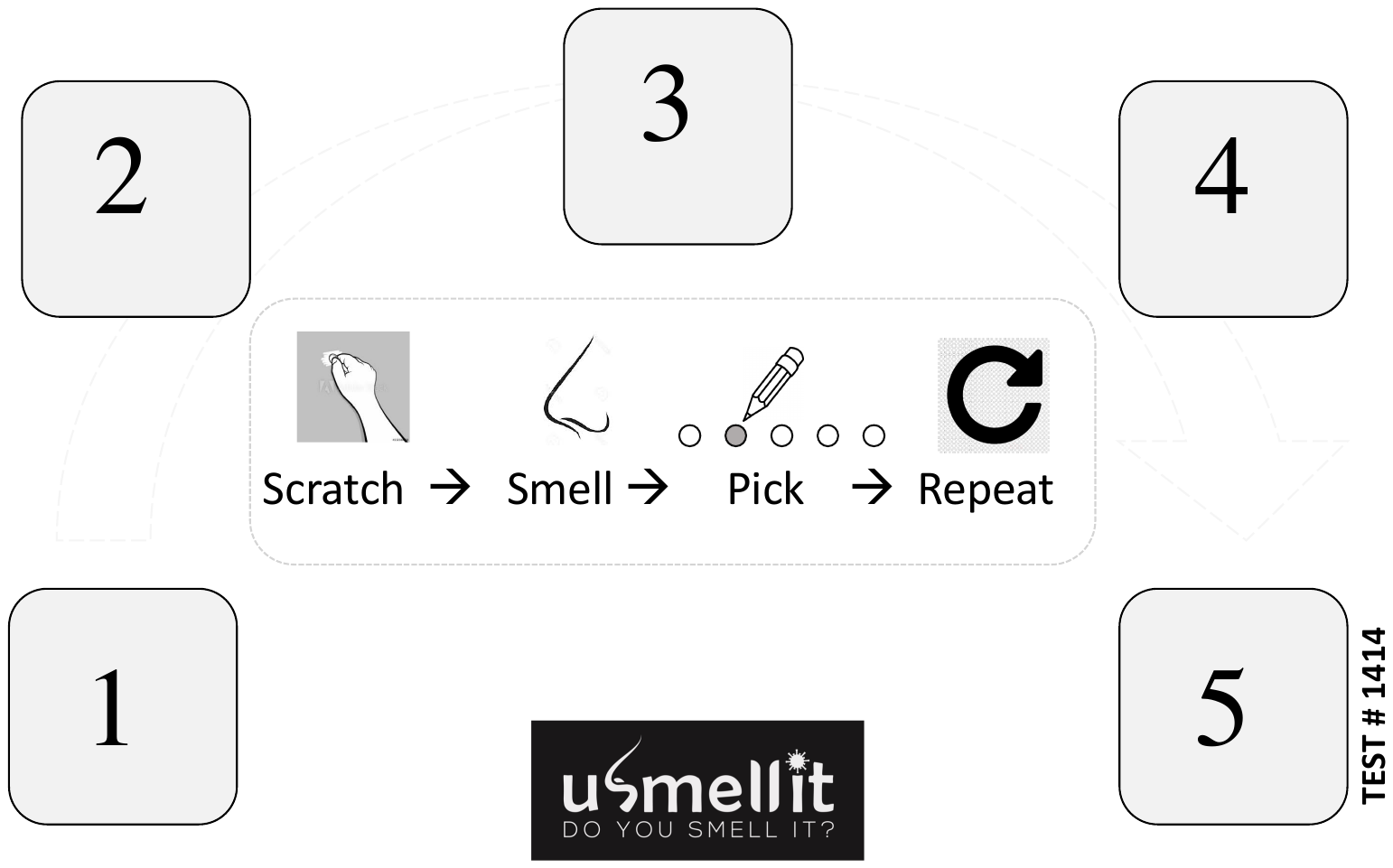

Supplemental Figure 2. Example of a response card to the olfactory test.

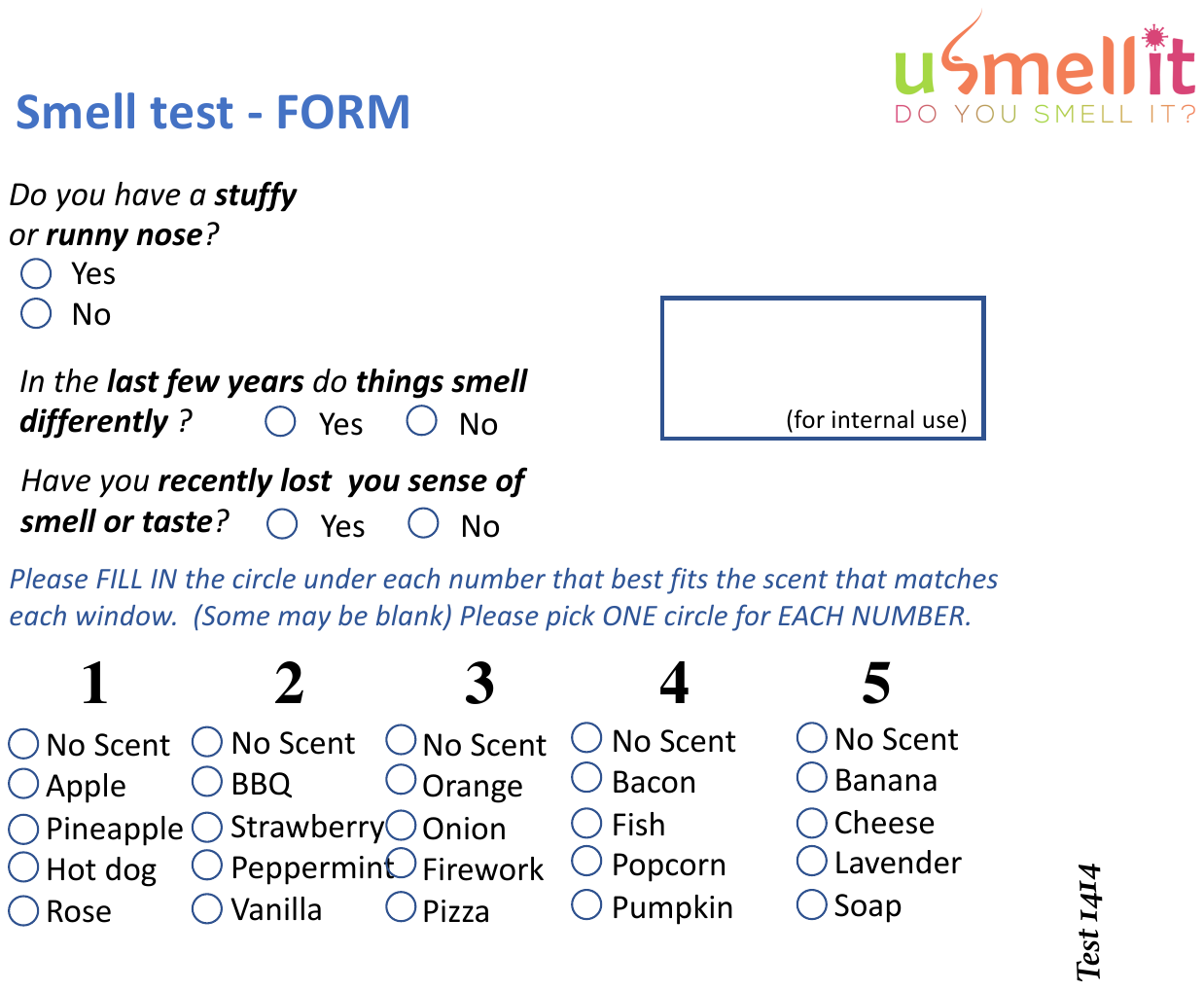

Supplemental Table 1. Scent options and the right response for all cards used in this study.

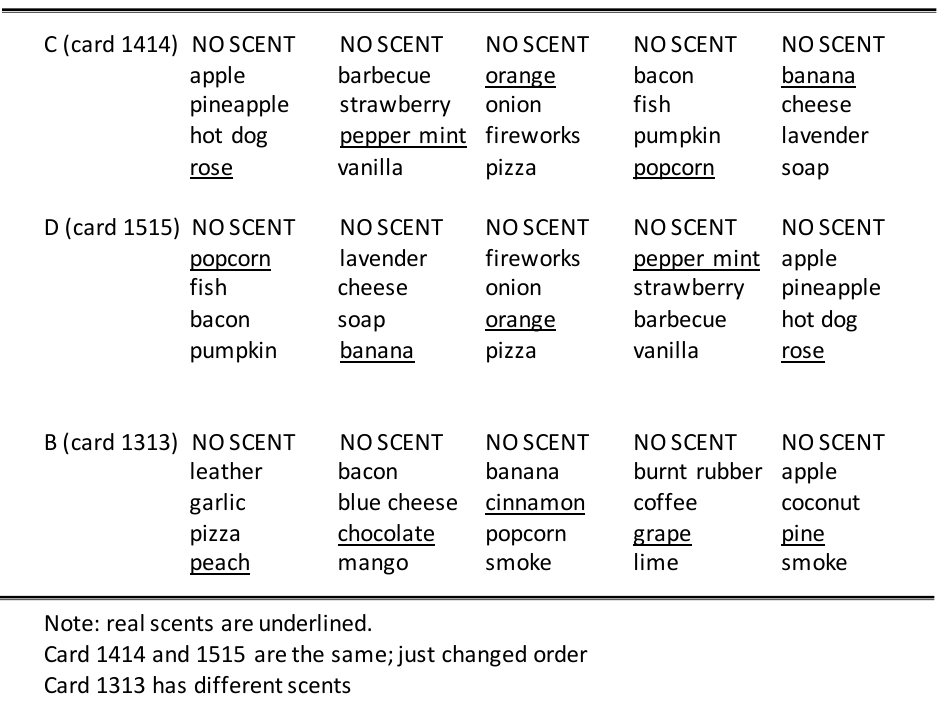
